# Second-generation mindfulness-based interventions (SG-MBIs) for university students: a scoping review

**DOI:** 10.1101/2025.10.20.25338430

**Authors:** Matthew A Aldridge, Nandacara, Alan Simpson, Karen O’Brien-Kop, Wasantha Priyadarshana

**Affiliations:** Florence Nightingale Faculty of Nursing, Midwifery & Palliative Care, King’s College London, London, United Kingdom; Postgraduate Institute of Pali and Buddhist Studies, University of Kelaniya, Bulugaha Junction, Kelaniya, Sri Lanka; Department of Health Service & Population Research, Institute of Psychiatry, Psychology and Neuroscience, King’s College London, London, United Kingdom; Department of Theology & Religious Studies, Faculty of Arts & Humanities, King’s College London, London, United Kingdom; Faculty of Buddhist Studies, Nāgānanda International Institute for Buddhist Studies, Manelwatta, Bollegala, Kelaniya, Sri Lanka

## Abstract

Second-generation mindfulness-based interventions (SG-MBIs) extend typical mindfulness-based interventions (MBIs) by grounding them in Buddhist epistemological and pedagogical frameworks and integrating additional Buddhist-derived practices. Systematic reviews show that MBIs can reduce burnout, distress, anxiety, depression and stress among university students—an issue of increasing global health concern—yet no reviews have specifically examined SG-MBIs in this population.

We conducted a scoping review following Joanna Briggs Institute guidance and reported in line with PRISMA-ScR. Prior to searches the protocol was registered with the Open Science Framework (https://osf.io/fyuqx/). Eight electronic databases and grey literature sources were searched for SG-MBIs delivered to university students between 2010 and 2025. Eligible studies included qualitative, quantitative and mixed-methods designs. Two reviewers, an English nursing researcher in the UK and a scholar-monk from Myanmar currently based at Kelaniya university in Sri Lanka, independently screened studies, extracted data and synthesized findings.

Sixteen publications describing eleven unique SG-MBIs were identified. Across the studies, 649 university students participated in SG-MBI interventions. Most interventions were delivered in high-income settings, with demographically homogeneous samples, narrow psychological outcome focus and predominantly expert-led models. SG-MBIs appeared safe and potentially effective as holistic health-promotion interventions within higher education. However, no studies reported integration with traditional healing systems, few assessed physical, social or spiritual well-being, and tailoring or personalisation were almost entirely absent. This should be addressed in future studies.

The Buddhist grounding of SG-MBIs suggests relevance for culturally congruent, scalable interventions, particularly in Buddhist-majority low- and middle-income countries. Future research should prioritize collaboration between researchers in high-income countries and specialists in Buddhist-majority LMICs to support development of culturally appropriate and sustainable interventions.

## Introduction

### Rationale

#### Student mental health and well-being

In an international survey of 14,000 students across 19 universities in 8 countries, 35% of students met the diagnostic criteria for at least one common mental health condition (1). University students reported higher levels of depression than the 12.9% reported in the general population and age matched peers (1,2). In the UK, the mental health and well-being of university students has become an important public health concern, the number of students saying they have a mental health condition increased sevenfold in the decade to 2020/21, up from 0.8% in 2010/11 (3–5). The promotion of musculoskeletal and mental health are key components of the WHO Europe Action Plan for non-communicable diseases (NCDs) (6) and Universities represent an important setting for health promotion (7–11).

#### Second-generation mindfulness-based interventions (SG-MBIs)

In a student mental health research priorities setting exercise, UK university students suggested the potential benefits of a broad range of specific and sometimes novel interventions including mindfulness-based interventions (12). Systematic reviews and meta-analyses have found that mindfulness-based interventions (MBIs) for university students can result in a significant reduction in burnout, distress, anxiety, depression and stress (13–18).

There is an emerging evidence-base for second-generation mindfulness-based interventions (SG-MBIs) (19,20) compared with first-generation mindfulness-based interventions (FG-MBIs) (21,22). However, systematic reviews and meta-analyses have been almost exclusively limited to FG-MBIs such as mindfulness-based stress reduction (MBSR) (23–25), mindfulness-based cognitive therapy (MBCT) (26) and adapted FG-MBIs. For example, one systematic review was limited to MBSR (18). Another review (15) was based on a previously proposed definition of mindfulness-based programmes (MBPs) (27), and acknowledged the analysis was limited to ‘…what have retrospectively become termed ‘first-generation’ MBPs –i.e. MBSR and MBCT… and the range of programs which have developed out of these…’ (27).

Both FG-MBIs and SG-MBIs are derived from Buddhist meditative practices (28), but key characteristics distinguish SG-MBIs from FG-MBIs (table 1). SG-MBIs ‘…are openly spiritual in nature, make the linkage to the Buddhist teachings explicit within the pedagogy and are more traditional in the manner in which they construct and teach mindfulness… These programs are at an earlier stage of development and research is now underway.’ (27).

**Table 1.**
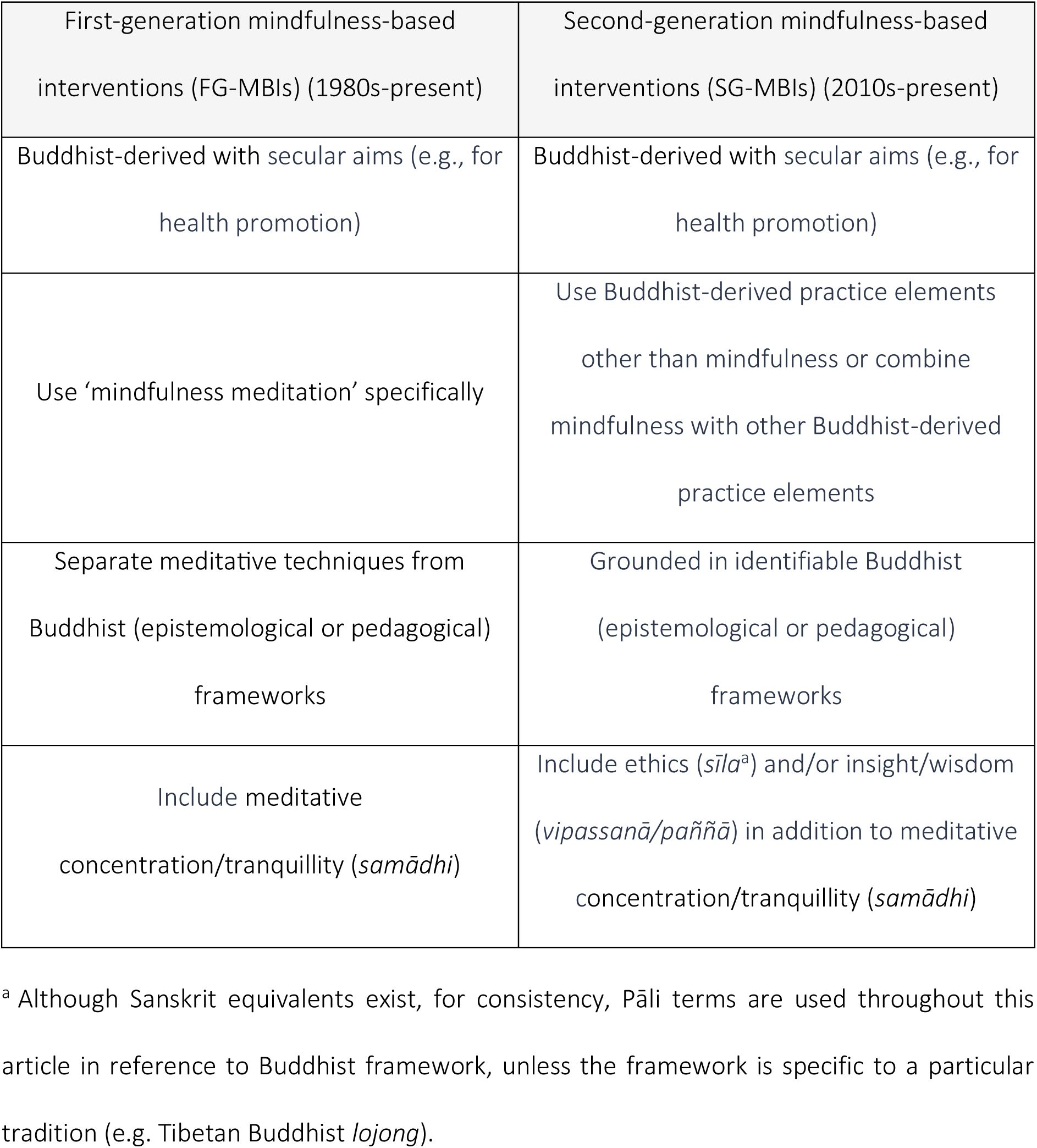
Features distinguishing second-generation from first-generation mindfulness-based interventions.

Whilst FG-MBIs have attempted to reductionistically extract meditative concentration (*samādhi*) from the threefold training (*tisikkhā*), from a Buddhist perspective no aspect is dispensable. The *tisikkhā* operates synergistically, and SG-MBIs were introduced to ‘…place mindfulness back within the *tisikkhā* structure of traditional Buddhist teaching’ (29–33). The *tisikkhā* includes:

1. Ethics/behaviour-change (*sīla*)
2. Concentration and tranquillity (*samādhi*)
3. Wisdom (*paññā*)

It has been argued that the *tisikkhā* must be present in any practice that claims to expound or be grounded in ‘authentic *Buddhadharma* [teachings of the Buddha]’ (34), and that it ‘…can be useful even for those who follow no religious tradition’ (35). It has important therapeutic applications (34,36–40) and the *Visudhimagga* (a classic manual of Buddhist doctrine and meditation) contains numerous ‘psychotherapeutic strategies’ centred around the *tisikkhā* (36). The *tisikkhā* has been applied as a ‘therapeutic system’ and used to explain Buddhist counselling processes wherein different aspects are prioritised at different times based on client conditions and needs (37,39,41).

Although current data is limited and preliminary, head-to-head randomised controlled trials (RCTs) comparing FG-MBIs with SG-MBIs have started to emerge and preliminary indications suggest SG-MBIs may have particular advantages (21,22,42,43), but until this review there have been no specific reviews of SG-MBIs for university students.

#### SG-MBIs for university students in Buddhist-majority South and Southeast Asian LMICs

Resources for, and research into the mental health of students in Buddhist-majority South and Southeast Asian low- and middle-income countries (LMICs) is limited (44–51). In Sri Lanka, for example, health promotion is crucial and calls continue to be made for health promotion interventions for university students (48,52–54).

There is evidence to support the feasibility and efficacy of MBIs for depression and anxiety in Asian countries (55,56) and when university students in Sri Lanka were asked to rate the helpfulness of a list of potential providers, clergy/religious priests were highly endorsed (57).

With their application of Buddhist frameworks, SG-MBIs could have potential to support the delivery of culturally acceptable and relevant health promotion for university students and others within Buddhist-majority countries, particularly LMICs such as Sri Lanka (58,59). Sri Lanka is facing multiple challenges in combatting non-communicable diseases (NCDs) and health promotion is crucial (53,54). Universities represent an important setting for health promotion (7–11) and calls have been made for mental health promotion interventions for university students in Sri Lanka where only a small proportion recognize their mental health difficulties and seek help (48,52).

#### Traditional, complementary, and integrative medicine (TCIM)

Use of traditional medicine in healthcare has been acknowledged in 88% of WHO Member States (60,61). Traditional healers are often the first point of contact for people with mental health conditions, who see their input as meaningful and likely to be helpful (62). Use of traditional healing for mental illness is widespread in many low- and middle-income countries (LMICs) as well as among marginalised populations within high income countries (HICs) (63). Traditional healers provide a substantial resource for mental healthcare where psychiatric treatment is lacking (64,65) and there are now ever increasing calls for collaboration between traditional and allopathic (biomedical) healing for mental disorders (65). The mental health workforce needs to be widened to integrate new ways of working with people, communities and organisations (66). There are likely many occasions where fruitful collaboration between health services and local healers is possible, and traditional practitioners, faith-based organizations and others have a crucial part to play within global mental health (62). Ayurvedic daily and seasonal routines (Sanskrit: *dinacaryā* and *ṛtucaryā*), for example, are claimed to have great potential for public health promotion (67–69) and have been advocated for the purpose of health promotion outside of Asia including for non-Asians (70,71).

A quasi-experimental study in Brazil found that daily Ayurvedic practices demonstrated ‘the potential to enhance quality of life’ and ‘contribute to health promotion’ in a ‘…practical and economically accessible manner’ (72) and a RCT in Southern California with 124 healthy adults assigned to either an Ayurvedic health promotion intervention, a ‘Western’ health promotion intervention, or control group, showed statistically significant improvements in both groups compared to control (73). On these grounds, the integration of SG-MBIs with traditional healing approaches such as Ayurveda may offer an important avenue for exploration.

#### Holistic Well-being

The World Health Organisation (WHO) defines health as ‘…complete physical, mental and social well-being’ (74). In relation to physical well-being there is significant scope for the implementation of lifestyle interventions to improve the health of university students (75,76). There is evidence supporting the potential of physical activity interventions in enhancing students mental health (77) and MBIs for adults in nonclinical settings which include physical exercise have been found to be more effective than MBIs without additional components (78). There is a significant correlation between unhealthy diet and increased risk of depression, anxiety and stress experienced by students (79,80). Addressing nutritional issues among university students has been identified as crucial for promoting good health and academic performance (81). Nutrition education may improve university students’ dietary intake (82) and the ‘NutriMind’ project provides an example of an intervention combining mindfulness with education on healthy diet for the management of depressive symptoms in university students within a LMIC (Uganda) (83).

Research suggests that acts of ‘giving’ and ‘kindness’ can improve well-being (84) and Berryman (85) argued that since moral influences on social well-being are well established (86–88), prioritizing ‘moral health’ (89), based on a person’s own sense of integrity in relation to their values, could be an important factor when considering the introduction of MBIs as a public health initiative (85).

According to Buddhist traditions, as well as having implications for contemporary behaviour modification and mental health (90), ‘recognising and transforming destructive emotions’ (such as greed or ill-will) lies at the heart of spiritual practice. Indeed: ‘…some hold that whatever lessens destructive emotions *is* spiritual practice’ (91). SG-MBIs have the potential to offer a holistic approach, addressing physical, mental, social and spiritual well-being.

#### Transdiagnostic Focus

Evidence supports the use of transdiagnostic interventions for common mental disorders (depression and anxiety symptoms) in primary care (92) and university-wide transdiagnostic mental health promotion interventions that target comprehensive skills training are likely to have greater reach than disorder-focused approaches and to fit well within university settings (11,93).

#### Precision/personalization/tailoring

Within transdiagnostic interventions a distinction has been proposed between “one size fits all” unified, and “my size fits me” individualized interventions which may be more consistent with a precision health approach (94,95). The case has been made for better understanding of which transdiagnostic factors predict the best MBI outcomes (96). It is important to examine for whom MBIs are most effective and for whom these interventions may not be effective (97). It has been suggested that greater knowledge of baseline moderators may optimize outcomes by better matching of individual participant characteristics to specific MBIs (97), and it has been recognised as important for SG-MBIs ‘…to examine moderators to see whether we can get at specificity and eventual treatment matching’ (97). Assessing participants’ personality traits has been recommended to enable ‘…a more tailored and beneficial approach’ to MBIs (98) and it has been proposed that the psychometrically validated Behavioral Tendencies Questionnaire (BTQ; 96,97): ‘…would seem to have potential implications for individualizing meditation and mindfulness-based practices.’ (99).

#### Delivery by Non-Specialist Providers (NSPs)

Systematic review and meta-analysis also supports transdiagnostic psychological interventions delivered by non-specialist providers (NSPs) in LMICs, for reducing symptoms of distress, anxiety, depression, and trauma (101) and models of NSP delivered health promotion may support communities in playing a stronger role in advocacy for health. However, a suggested aspect of SG-MBI delivery has been the use of an instructor training program that ‘…normally requires several years of supervised mindfulness practice’ (19,20) the extent to which this would limit delivery by NSPs is an important consideration potentially limiting SG-MBI potential in this respect.

### Aim and Objectives

The scoping review followed Joanna Briggs Institute (JBI) guidance (102), which builds upon earlier methodological frameworks for scoping studies proposed by Arksey and O’Malley (103) and further refined by Levac et al. (104). The review aimed to answer the primary research question:

“What second-generation mindfulness-based interventions (SG-MBIs) have been delivered with university students?”

Within each identified SG-MBI, the objectives were to address secondary research questions (see S1 Appendix) relating to:

- Geographical context
- Participant demographics
- Buddhist epistemological or pedagogical frameworks and practice elements
- Integration of traditional medicine
- Approaches to well-being
- Tailoring
- Delivery models

## Methods

### Study design

This scoping review was undertaken in accordance with the Preferred Reporting Items for Systematic Review and Meta-Analyses Extension for Scoping Reviews (PRISMA-ScR) (99); see S2 Appendix. Prior to searches the protocol was registered with the Open Science Framework (https://osf.io/fyuqx/). The review was steered by a team including multidisciplinary experts in Sri Lanka and the UK who contributed to the research questions.

### Eligibility criteria

#### Population

University students (undergraduate and postgraduate) aged 18 or over, of any gender, ethnicity, sexuality, etc.

#### Setting

We set out to include studies that focused (or provided disaggregated data) on SG-MBIs delivered within university settings (higher education institutions globally), and other settings where SG-MBIs were used, and university students (at least one) included as participants.

#### Intervention

For the purpose of this scoping review, drawing on previous conceptualisations (19,20,27,41), SG-MBIs were conceptualised on the basis of meeting the criteria outlined in table 1. Interventions which incorporated Buddhist-derived practice elements such as appreciative joy meditation (AJM) (101,102), without grounding within a Buddhist epistemological or pedagogical framework, were excluded. See S3 Appendix for full inclusion/exclusion criteria.

#### Outcomes

We included studies that reported any positive or adverse outcomes of SG-MBIs, whether from implementation or experimental evaluation. No restrictions were placed on the type of outcomes assessed. Consistent with PRISMA-ScR guidance, we did not synthesise effect sizes or effectiveness data across studies. Instead, we charted and thematically mapped the outcome measures employed in relation to four well-being domains (physical, mental, social, and spiritual) to provide an overview of how SG-MBIs with university students have been evaluated.

#### Types of Studies

We included quantitative, qualitative or mixed-methods empirical research study designs. Both peer reviewed and grey literature sources (English only) were eligible providing they included a description of methodology employed. PhD theses were included unless the same study was later peer-reviewed and published, in which case the published version was used instead.

### Search strategy

A three-step search strategy was used. Firstly, between 11^th^ July 2025 and 8^th^ August 2025, we searched five databases: EMBASE; MEDLINE; PsychINFO; Global Health; AMED. An example full search strategy is available in S4 Appendix. Searches were also run in two electronic grey literature databases (Global Health and Google Scholar), two pre-print servers (medRxiv and PsyArXiv), and one PhD thesis website (Open Access Theses and Dissertations; OATD). The search strategy began with using terms adapted from editorial articles on SG-MBIs (19,20) and related systematic reviews with a focus on FG-MBIs for university students and on Buddhist-derived loving-kindness-, compassion- and wisdom-based meditation practices (13–18,98,107). Since SG-MBIs have been emerging only since the mid-2010s searches were limited to works post 2010 in English language. Secondly, forward citation searching was conducted using Web of Science and Google Scholar for all studies meeting inclusion criteria. Reference lists of all included studies were checked for relevant studies. Finally, international experts were contacted to identify additional studies. See figure 1 for PRISMA diagram demonstrating search strategy.

**Fig 1.**
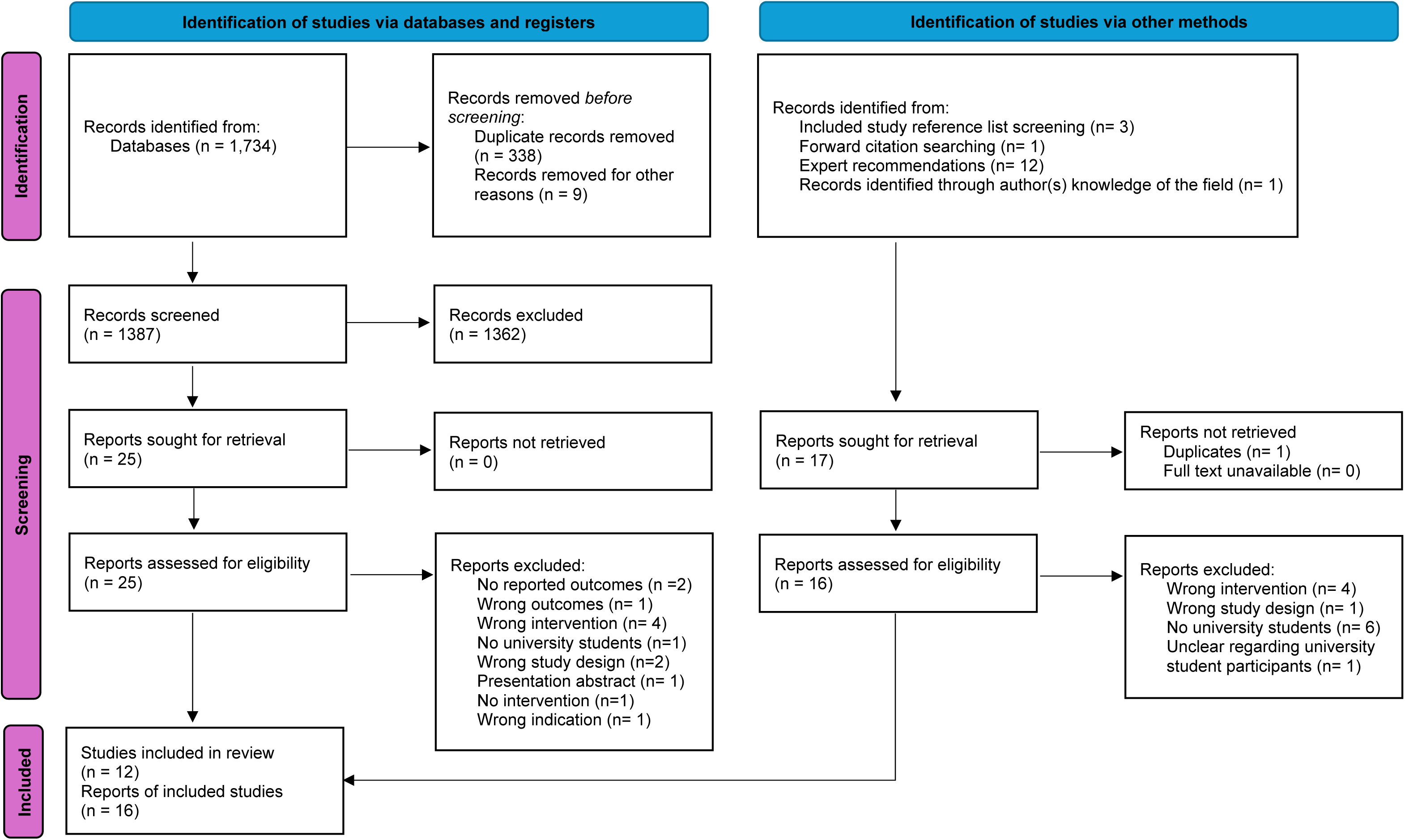
PRISMA diagram demonstrating search strategy (109)

#### Study selection

All studies identified through database searches were independently screened by an English nursing researcher in the UK (MA) and a scholar-monk from Myanmar who is currently based at Kelaniya university in Sri Lanka (NC), with disagreements resolved through discussion. Screening was conducted via Covidence (108). Studies identified through forwards and backwards citation searching and expert recommendation were also double screened independently by MA and NC.

#### Charting and organising

A data extraction form template based on the research questions was developed by MA using Covidence and revised collaboratively with NC. The form was piloted and refined as needed to ensure all relevant data were captured consistently. Data extraction was completed by MA and NC and double extracted to ensure accuracy.

### Data synthesis process

Data relevant for each research question were synthesised narratively by MA and NC. Question 1 entailed independent use by MA and NC of the four criteria above (table 1) by which interventions were agreed to be SG-MBIs. Where data were available, evidence for questions 2-10 (S1 Appendix) was synthesised within each SG-MBI. Data were narratively synthesised together for each question through discussion between MA and NC. To address question 6 on whether the SG-MBI promotes a holistic approach to well-being, outcome measures reported in each study were independently reviewed and mapped to four well-being domains—physical, mental, social, and spiritual—by the review team. Where measures had potential overlap between domains, assignment was determined by consensus between MA and NC. This process ensured consistency while acknowledging the subjective nature of domain classification.

## Results

### Study Characteristics

The included research papers were published between 2013 and 2025. Most studies were RCTs (n= 9), but studies also included a controlled pilot study, an uncontrolled trial, a mixed-methods design, a pilot study using case studies and a qualitative analysis of service-user experiences. All, except one PhD thesis, were articles published in peer-reviewed journals. See table 2 for characteristics of all included studies.

**Table 2.**
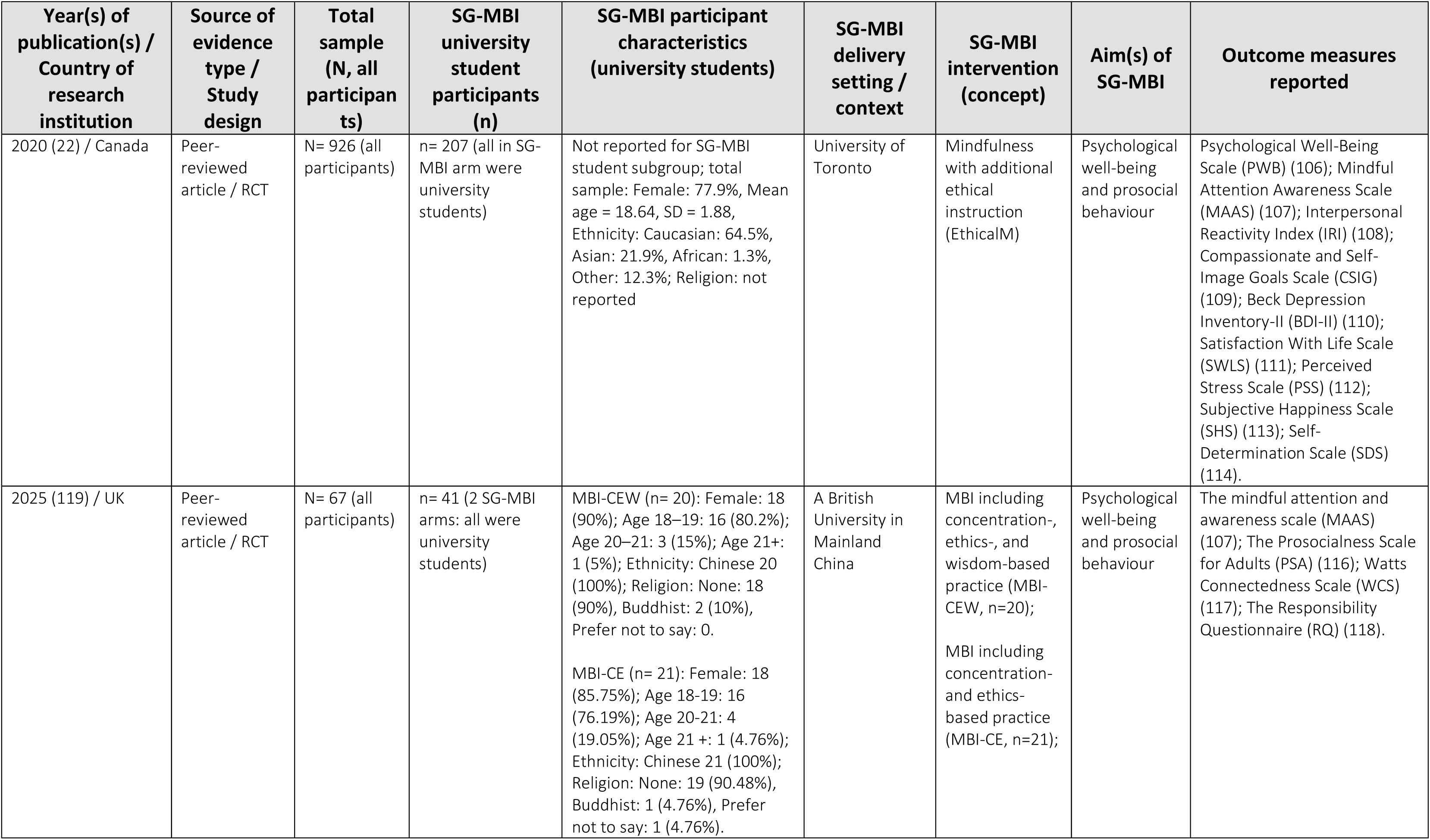
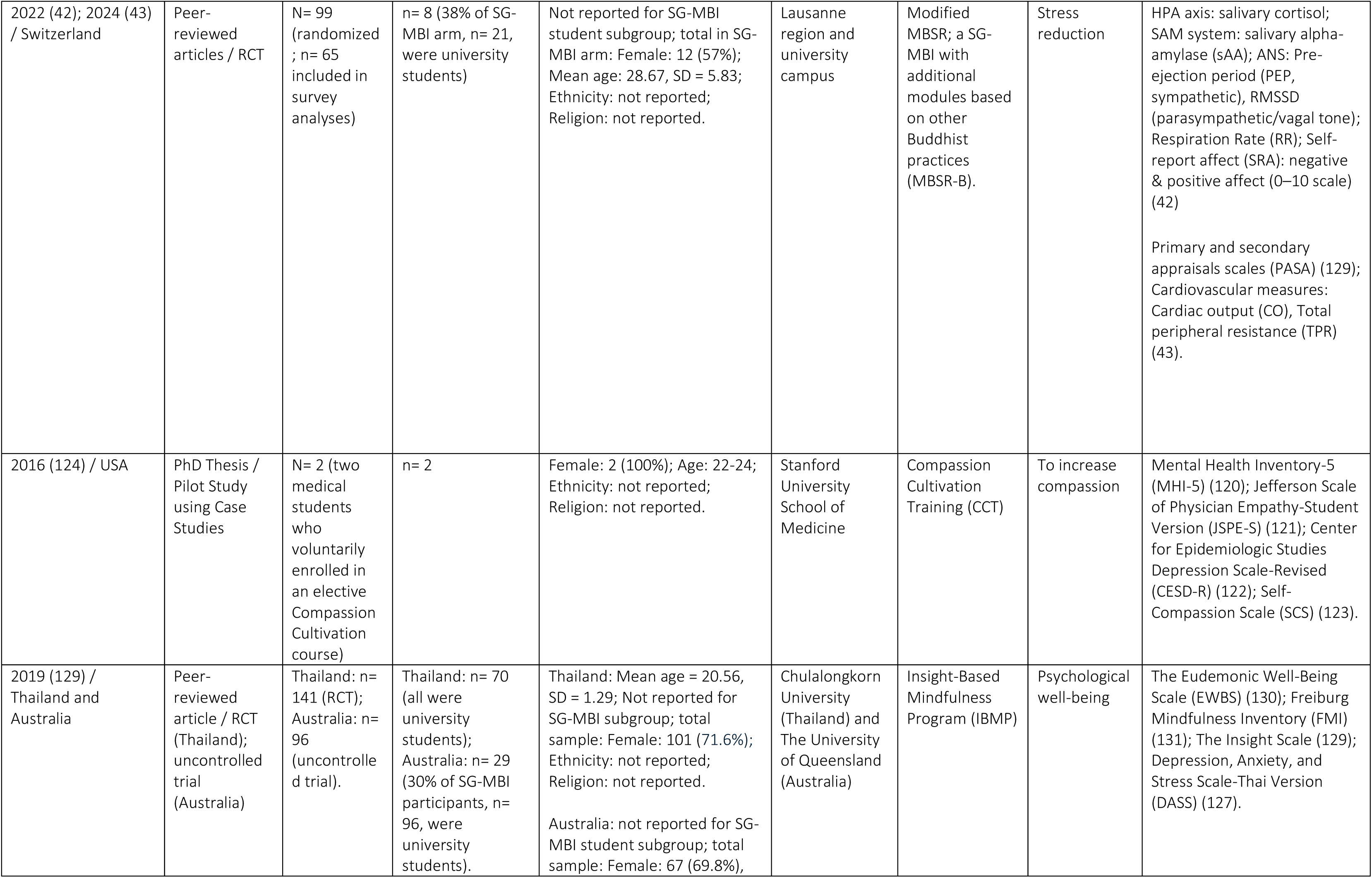
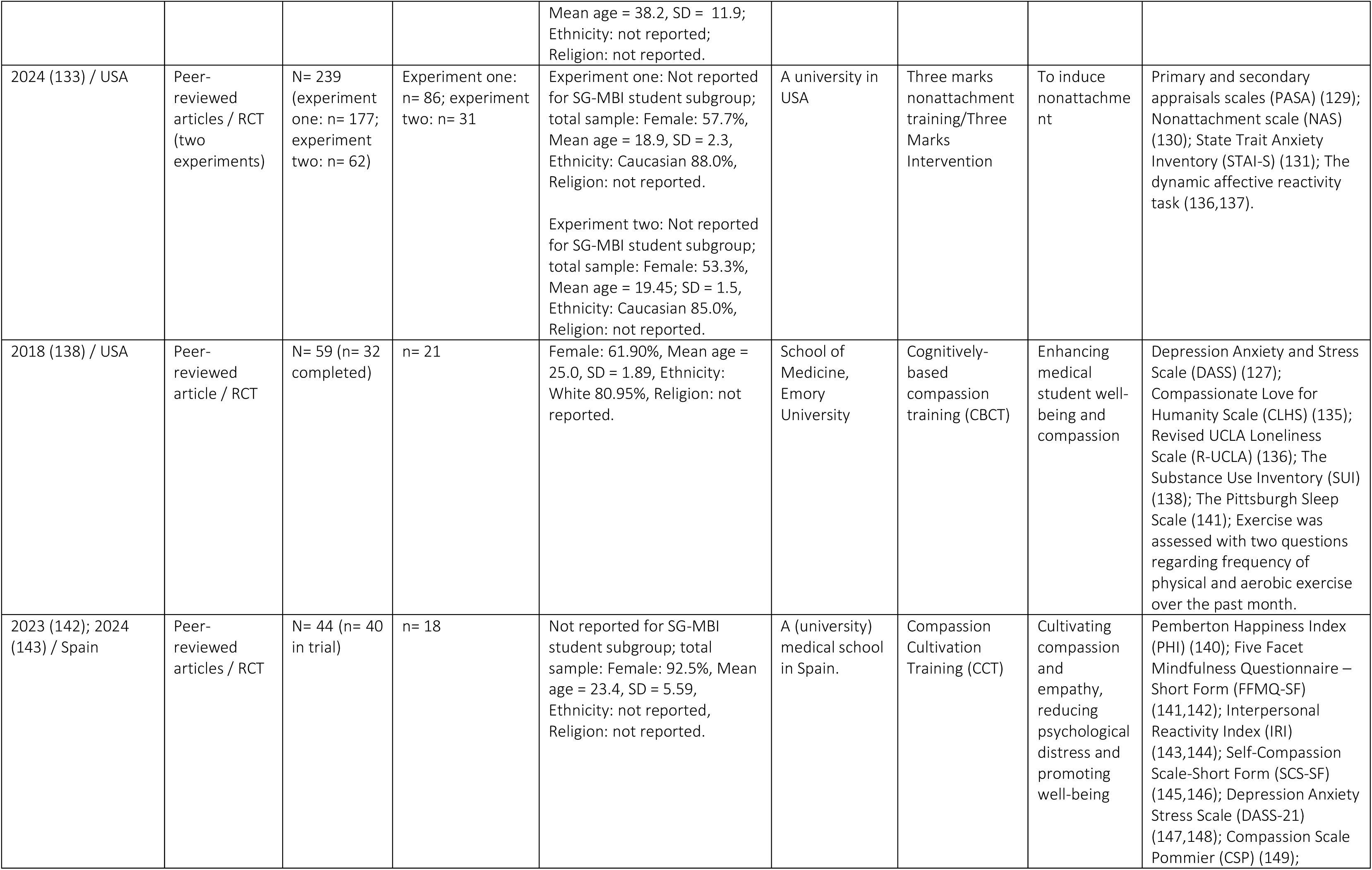
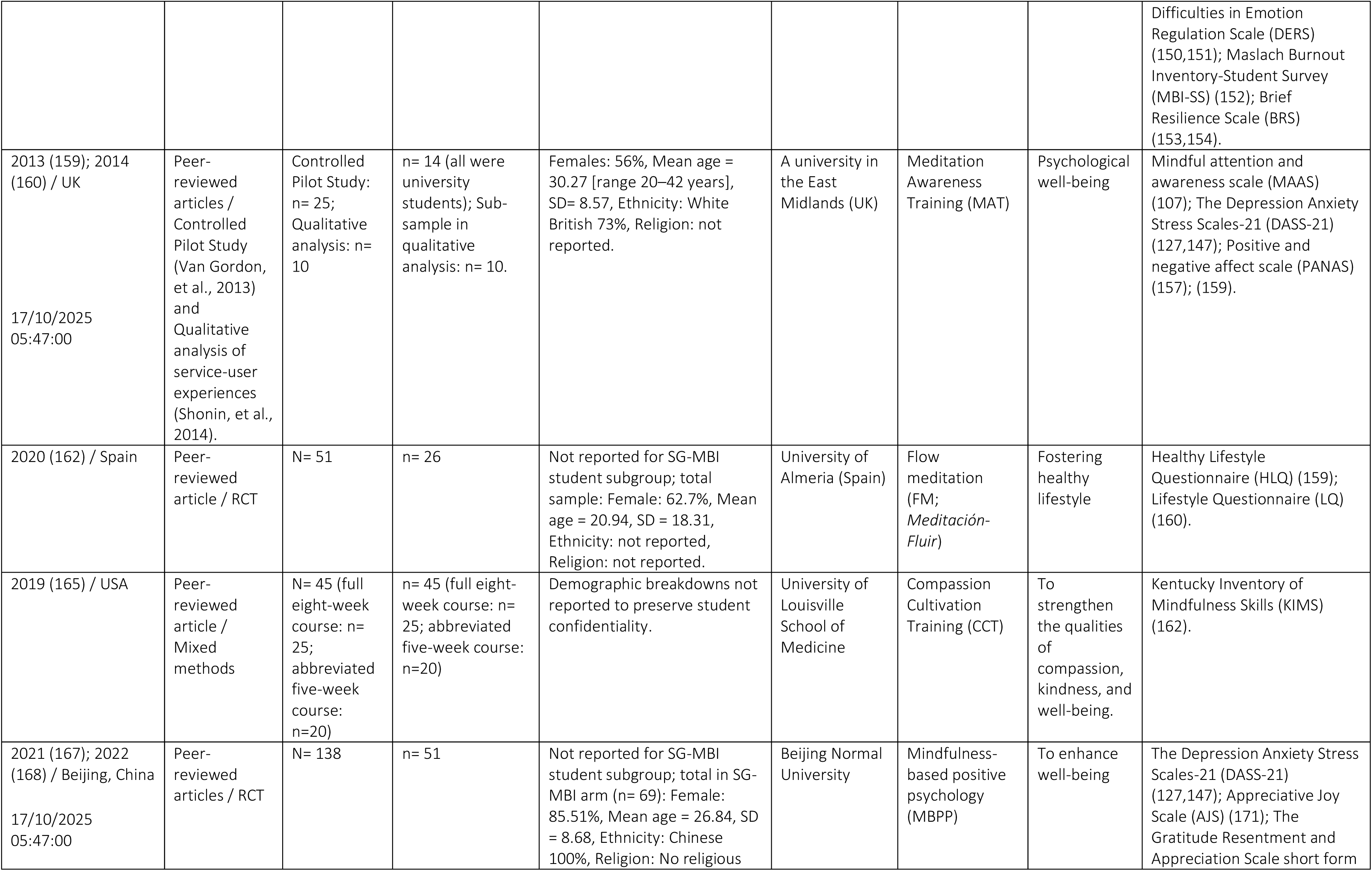
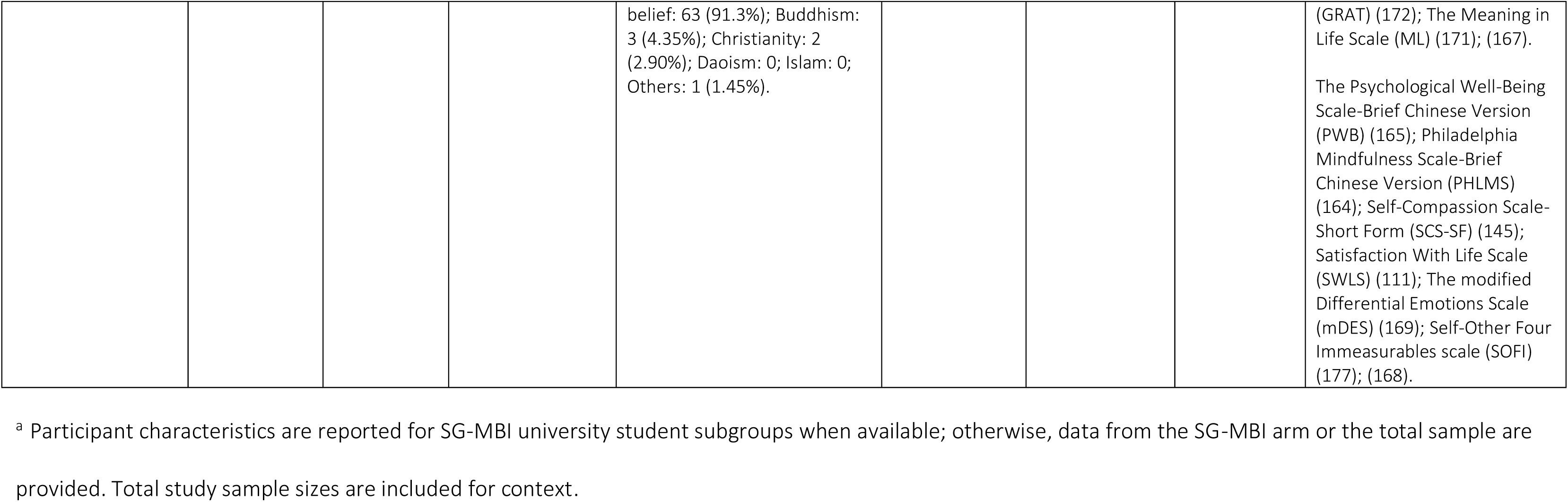
Characteristics of included sources of evidence^a^.

Although spanning a wide geographical range including North America (USA, Canada), Europe (UK, Switzerland, Spain), Asia (China, Thailand), and Australia, the studies were all conducted in high-income countries (HICs), except for one RCT which included 141 university students in Thailand.

### Population Details

The sixteen included publications (15 peer-reviewed articles and one PhD thesis) reported on 12 intervention studies. Across these studies, a total of 649 university students participated in SG-MBIs, with individual study sample sizes ranging from a very small pilot (n = 2) (119) to large-scale RCTs (e.g., n = 926) (22). All studies had female-majority samples, ranging from 53.3% to 100%. Mean ages ranged from approximately 18.6 to 28.7 years. Half of the included publications (n = 8/16) did not report participant ethnicity and among those that did, participants in North America (USA, Canada) and the UK were predominantly White/Caucasian, while participants in China were entirely Chinese. Religious affiliation was reported in only a few of the publications (n = 4/16). Where recorded, most students identified as having no religion, with only small minorities identifying as Buddhist or Christian.

### Buddhist Frameworks and Practice Elements

11 SG-MBIs were identified within this scoping review:

1. Cognitively-Based Compassion Training (CBCT) (134)
2. Compassion Cultivation Training (CCT) (124,142,143,165)
3. EthicalM (22)
4. Flow Meditation (FM) (162)
5. Insight-Based Mindfulness Program (IBMP) (129)
6. Meditation Awareness Training (MAT) (159,160)
7. Mindfulness-Based Intervention including Concentration- and Ethics-based practices (MBI-CE) (119)
8. Mindfulness-Based Intervention including Concentration-, Ethics-, and Wisdom-based practices (MBI-CEW) (119)
9. Mindfulness-Based Positive Psychology (MBPP) (167,168)
10. Modified Mindfulness-Based Stress Reduction (MBSR-B) (42,43)
11. Three Marks Intervention (TMI) (133)

These SG-MBIs incorporated a range of Buddhist frameworks, most frequently the threefold training (*tisikkhā*) (n = 7/11), the three marks of existence (*tilakkhaṇa*) (n = 4) and the four immeasurables (*brahmavihārās*) (n = 2). Intervention content typically included practices relating to ethics/behaviour-change (*sīla*) (e.g., compassion, generosity, non-harming), concentration and tranquillity (*samādhī*) (e.g., mindfulness of breath, *shamatha* (*samatha*), and wisdom (*paññā*) (e.g., impermanence (*anicca*), interdependence and non-self (*anatta*)). As seen in table 3, while several interventions integrated all three domains, others focused on concentration (samādhī) and wisdom (*paññā*), or exclusively on wisdom. See S5 Appendix.

**Table 3.**
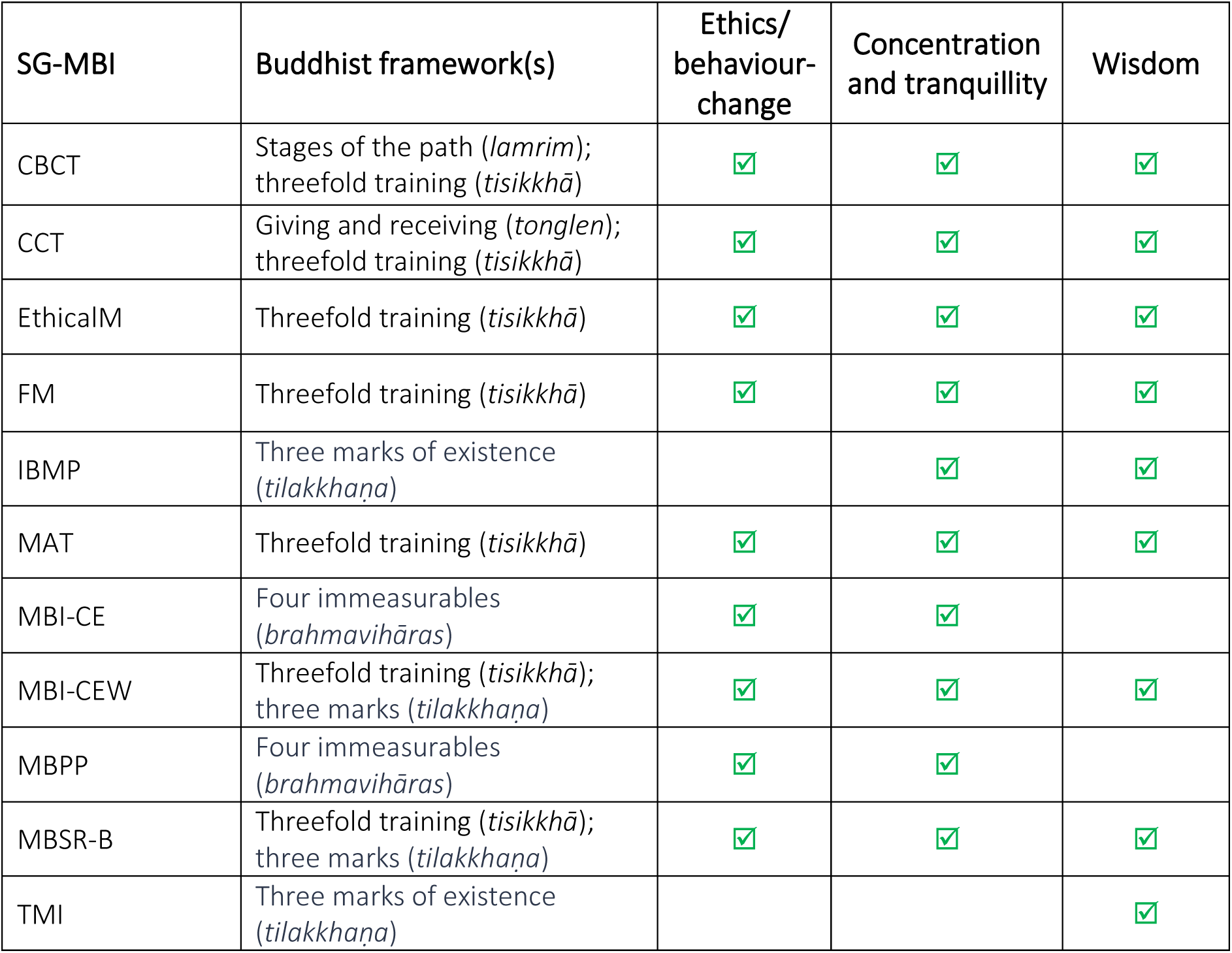
Buddhist Frameworks and Practice Elements.

### Traditional, complementary, and integrative medicine (TCIM)

None of the SG-MBIs incorporated aspects of Ayurveda, Traditional Thai Medicine (TTM) or other traditional healing elements.

### Holistic Well-being

Through consensus between reviewers, the outcome measures in each study were mapped to four domains of well-being (physical, mental, social, spiritual). SG-MBIs varied in their focus. With the exception of the RCT of Flow Meditation (FM; which has been described as one of the first MBIs to specifically investigate the effects of mindfulness on healthy lifestyle in university students (162)), aspects of mental well-being were assessed in all other SG-MBIs (n = 10/11). Aspects of social well-being were assessed in CCT, EthicalM, MBI-CE and MBI-CEW (n = 4/11). Aspects of physical well-being were assessed in FM, CBCT and MBSR-B (n = 3/11). Aspects of spiritual well-being were assessed in CBCT, CCT, IBMP, MBPP and TMI (n = 5/11). See S6 Appendix.

### Transdiagnostic Focus

None of the identified SG-MBIs had a disorder-specific focus; rather, they adopted a transdiagnostic approach, which has been identified as likely to have a greater reach and to fit well within university settings (11,93). The SG-MBIs broadly targeted psychological well-being through dispositional mindfulness, emotion regulation (e.g., neuroticism, compassion, empathy), stress processes (stress response, resilience to stress and burnout), lifestyle factors (healthy lifestyle behaviours) and broader well-being outcomes (life satisfaction, prosocial behaviour and self-actualisation).

### Precision/personalization/tailoring

Personalization within the SG-MBIs was very limited. One exception was a component of the MAT intervention, which entailed 50-minute one-to-one support sessions during weeks 3 and 7 to discuss individual progress, address training-related issues and provide personalized guidance (159). Aside from this component, the SG-MBIs generally took a “one size fits all” unified transdiagnostic approach (94,95).

### Delivery Models

Across the included studies, the dominant model for SG-MBI delivery was in-person group sessions supported by home practice using guided recordings, workbooks, or informal assignments (n = 9/12). A smaller number of studies used online classes/group sessions (n = 1/12) or pre-recorded audio/video delivery (n = 2/12). SG-MBI providers in the studies were typically certified instructors or highly experienced meditation teachers (n = 10/12). Compassion-based interventions (CCT, CBCT) and MBSR-B required formal teacher certification, while other interventions (FM, MBPP, IBMP, MAT) emphasized the requirement of extensive meditation practice, teaching experience and relevant knowledge. Only interventions centred on audio/video pre-recordings (TMI, EthicalM) appeared to demonstrate potential feasibility for non-specialist provider (NSP) delivery.

SG-MBI duration ranged from very brief to more extended formats lasting up to 10 weeks. The Three Marks Intervention (TMI) was a single 14-minute exposure and EthicalM entailed 6-days of 10-minute daily audio-guided practice. The other 9 SG-MBIs lasted between 4 and 10 weeks, with most interventions (n = 8) delivered in 7–9 weeks. Both online and in-person classes/group sessions were typically for 1.5-2.5 hours weekly, with home practice expectations ranging from 10 minutes daily (EthicalM) to 55 minutes daily (MBSR-B), most commonly between 20-40 minutes daily.

### Study Outcomes

The purpose of this scoping review was not analysis of effectiveness, feasibility, acceptability or participant engagement. No adverse outcomes were reported in any of the studies.

## Discussion

This scoping review identified eleven distinct SG-MBIs for university students, involving 649 participants. Most studies were conducted in high-income countries (HICs) samples were largely ethnically homogeneous and few studies reported religious affiliation. Considering the relevance and potential for SG-MBIs within Buddhist-majority LMICs this highlights serious issues around equity.

While nearly all studies measured mental well-being, fewer assessed physical, social, or spiritual well-being, suggesting evaluations remain psychology-focused rather than holistic. For population health more broadly, SG-MBIs should also focus on physical, social, and spiritual dimensions of well-being.

All SG-MBIs adopted a transdiagnostic approach, which aligns with university populations. Although evidence supports transdiagnostic interventions delivered by non-specialist providers (NSPs) in LMICs (101), most SG-MBIs relied on highly experienced meditation teachers. However, some interventions (e.g., TMI, EthicalM) that used pre-recorded content indicate potential feasibility for NSP delivery.

Despite recognition of tailored approaches as a future direction for SG-MBIs (97–99), personalisation was largely absent (with only MAT offering individualised guidance). No studies integrated traditional healing systems such as Ayurveda, despite potential cultural relevance, particularly in majority-Buddhist LMICs such as Sri Lanka and Myanmar.

Although effectiveness was not the review focus, no adverse outcomes were reported, supporting SG-MBIs as safe for students. Nonetheless, consistent reporting of harms should be prioritised in future research.

Moving forward, SG-MBI development should prioritise inclusive and culturally sensitive designs, integration with traditional health systems, and feasibility testing with NSPs. Partnerships between researchers in HICs and specialists in Buddhist-majority LMICs – such as those advancing Buddhist Ayurvedic Studies in Sri Lanka (36,38,39,178–188)– could play a critical role in shaping equitable and globally relevant health promotion strategies.

### Limitations

Whilst clear and precise criteria distinguishing SG-MBIs from FG-MBIs have been undergoing refinement for about a decade (19,20,189), the strict criteria applied in this review (table 1) may be a limitation. Only interventions grounded in Buddhist frameworks and explicitly addressing ethics (*sīla*) and/or wisdom (*paññā*), were included. Buddhist-derived interventions employing elements such as loving-kindness meditation (LKM; *metta*, 175) or appreciative joy meditation (AJM; *muditā*, 101,176), without grounding in such frameworks, were excluded. This may have narrowed scope, but preserved conceptual clarity of SG-MBIs as referring to interventions which ‘…make direct use of Buddhist principles’ (20) and ‘…make the linkage to the Buddhist teachings explicit within the pedagogy…’ (27).

A further limitation of this review is its potential contribution to the *‘*psychologisation’ of Buddhist frameworks (192). *Brahmavihāra* meditation (BVM) (also called the four immeasurables meditations; FIM) (193,194) were classified as ‘ethics-based practices’ within some of the SG-MBIs in this review (119,167,168). However, traditional sources such as the *Visuddhimagga* (ca. 400 CE) treat the *brahmavihārās* primarily as concentration (*samādhi*) rather than ethics (*sīla*) practices (195). Framing them as ethical practices risks undermining claims of SG-MBIs “authenticity” relative to traditional Buddhist frameworks (20,196). Nevertheless, some scholarship positions the *brahmavihārās* at the intersection of moral cultivation (*sīla*) and concentration (*samādhi*) (193), and empirical evidence supports their connection with ethical decision-making (197).

Informal practices within MBSR-B include ‘abstaining from one minor unethical action for the week (like gossiping), practicing one generous action…’ (42). So ethics-based practices within current SG-MBIs are not limited to contemplative exercises. Nevertheless, stricter criteria for classifying ethics-based practices (*sīla*) within SG-MBIs as including ethics-based behavioural change/regulation in relation to speech and action would be suitable towards ensuring consistent alignment with traditional frameworks.

## Conclusions

SG-MBIs have an important potential for health promotion among university students and for the public more broadly, and since they retain closer links to Buddhist frameworks than first-generation approaches, SG-MBIs may be particularly relevant for supporting health promotion within Buddhist-majority LMICs. However, the current evidence-base remains narrow in scope, most interventions were developed and delivered in high-income settings with limited diversity in samples, outcomes, and delivery models. SG-MBIs appear safe for students, but systematic harms reporting should continue to be prioritised.

Tailored transdiagnostic approaches, non-specialist provider (NSP) delivery models and the potential for integration with traditional healing systems require further exploration. Future development and trials of interventions that are grounded in Buddhist frameworks, or that integrate Buddhist-derived practices, should move towards ‘reciprocal innovation’ which involves ‘bidirectional, mutually beneficial research’ (198).

Global health reciprocal innovation (GHRI) describes a ‘…distinct, mutually beneficial and equitable approach to global health research that is focused on the transfer and exchange of health innovations around the world’ (199). In the development of interventions on the lines of SG-MBIs this should include collaboration between researchers in HICs and specialists in Buddhist-majority LMICs—such as those advancing Buddhist Ayurvedic Studies in Sri Lanka (36,38,39,178–188). These efforts are essential to realising the potential of SG-MBIs, as equitable and globally relevant public health interventions.

## Data Availability

All data supporting the findings of this study are included within the manuscript and its supplementary files.

## Acknowledgements

The primary author was originally supervised and mentored by the late Emeritus Prof. Sumanapala Galmangoda (1951–2024), a visionary scholar who pioneered and established Buddhist Ayurvedic Studies as a distinct applied sub-field within Buddhist Studies in Sri Lanka (38,39,180–184). With support from the WHO, Prof. Galmangoda founded this emerging discipline and championed the vision that Buddhist Ayurvedic Studies could gain recognition and contribute to global well-being. His guidance, insight, and dedication continue to inspire this work.

## S1 Appendix. Research Questions

The scoping review sought to answer the following primary research question:

1. What SG-MBIs have been delivered with university students?

Within each identified SG-MBI, the following secondary research questions were also addressed:

2. In which geographical regions was the SG-MBI developed, and what were the demographics (e.g., ethnicity, country, LMIC/HIC setting) of study participants?
3. What Buddhist pedagogical framework(s) were used within the SG-MBI?
4. What Buddhist-derived ethics/behaviour-change (*sīla*)-based, contemplation (*samādhi*)-based, and/or wisdom (*paññā*)-based practice elements were incorporated within the SG-MBI?
5. Did the SG-MBI integrate aspects of traditional medicine or healing?
6. Did the SG-MBI promote a holistic approach to well-being, addressing physical, mental, social, and spiritual dimensions?
7. Was the SG-MBIs disorder-specific (e.g., for depression) or transdiagnostic (e.g., targeting broader outcomes such as psychological well-being)?
8. Did the SG-MBI incorporate a precision/personalized/tailored approach?
9. Was the SG-MBI delivered by non-specialist providers (NSPs), or is there evidence supporting their feasibility for NSP-led delivery?
10. What study design (e.g., RCTs, qualitative, mixed-methods) was used to evaluate the SG-MBI for university students

## S2 Appendix. Preferred Reporting Items for Systematic reviews and Meta-Analyses extension for Scoping Reviews (PRISMA-ScR) Checklist

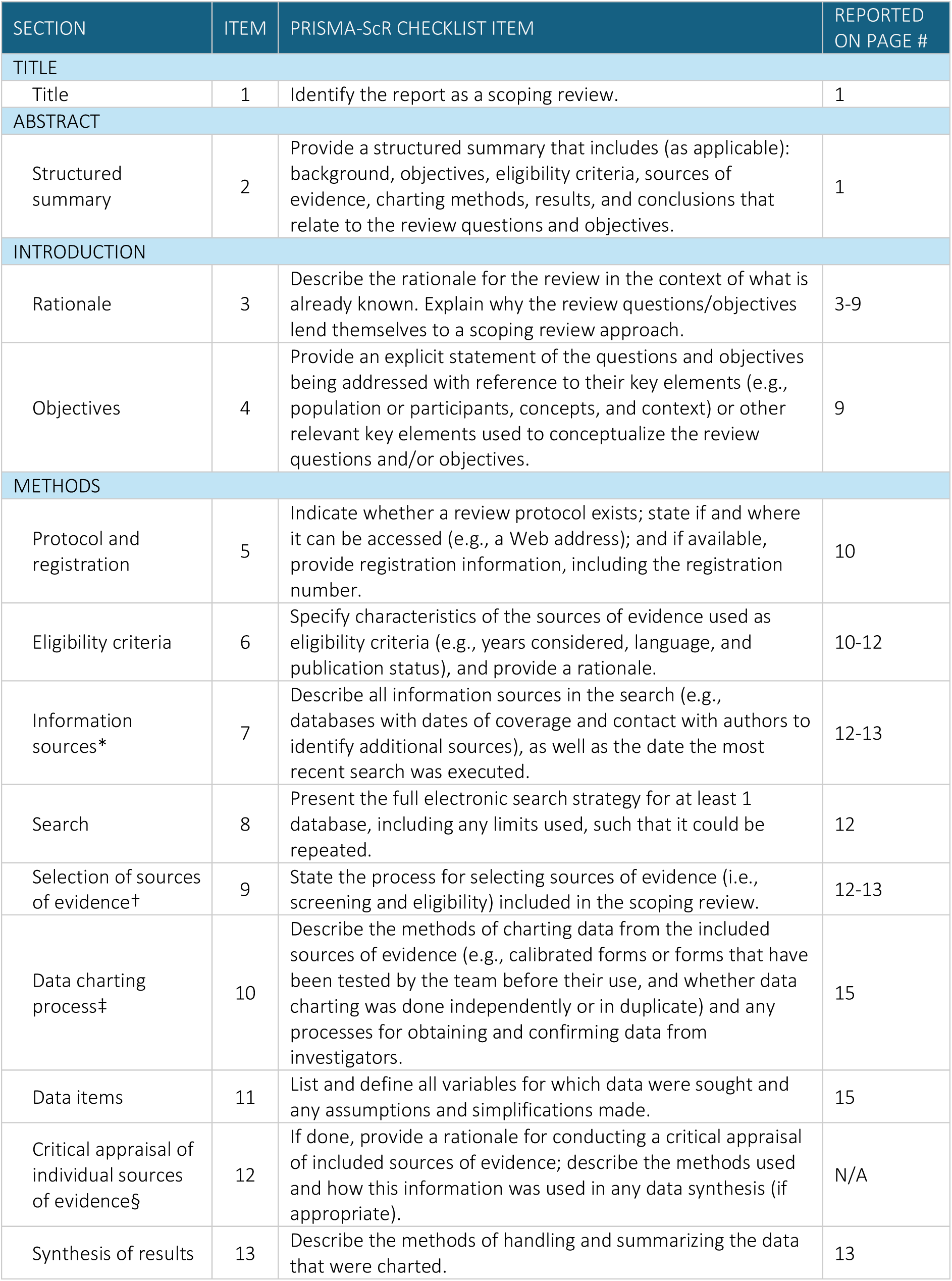
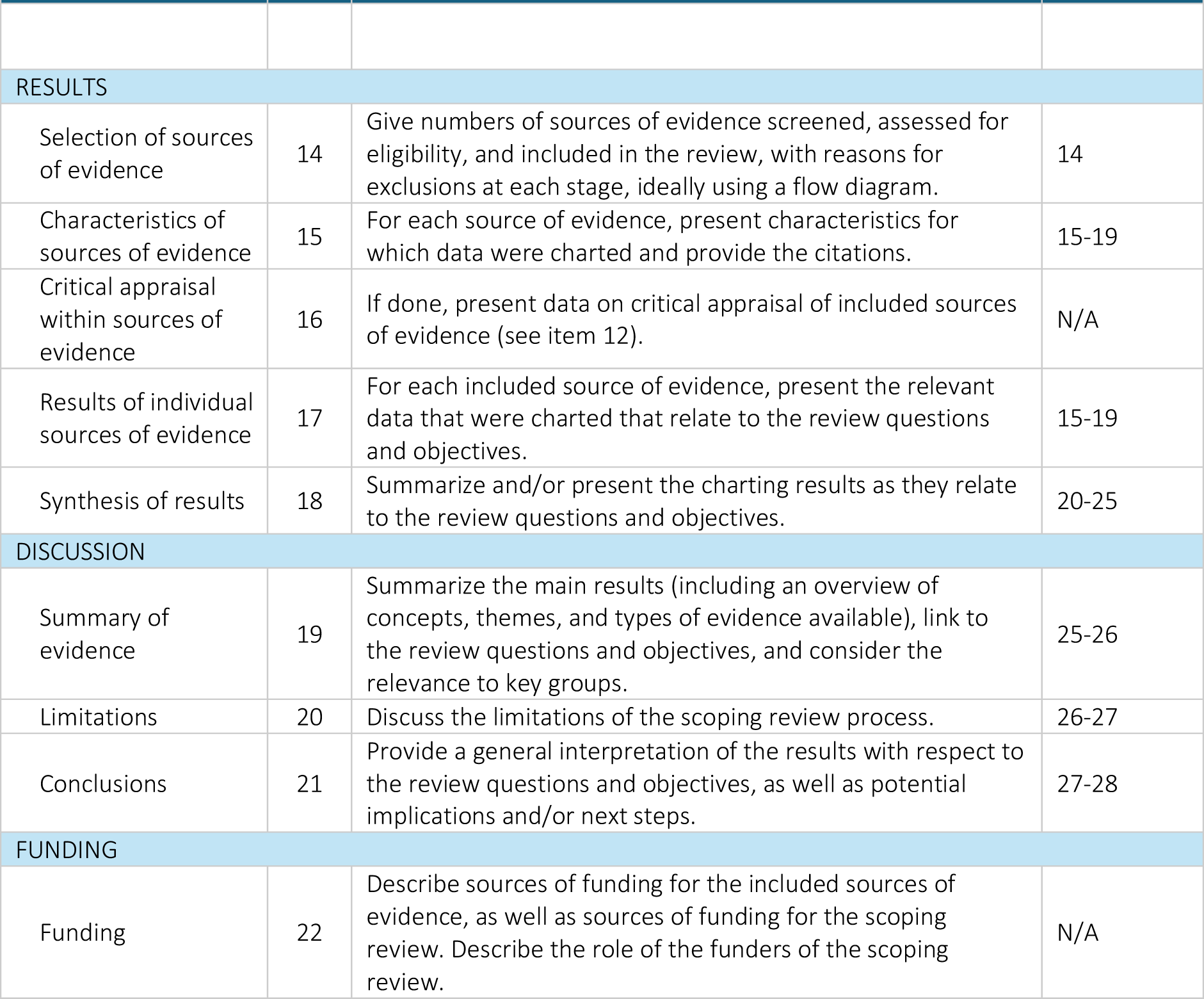

| SECTION | ITEM | PRISMA-ScR CHECKLIST ITEM | REPORTED ON PAGE # |
| --- | --- | --- | --- |
| TITLE |  |  |  |
| Title | 1 | Identify the report as a scoping review. | 1 |
| ABSTRACT |  |  |  |
| Structured summary | 2 | Provide a structured summary that includes (as applicable): background, objectives, eligibility criteria, sources of evidence, charting methods, results, and conclusions that relate to the review questions and objectives. | 1 |
| INTRODUCTION |  |  |  |
| Rationale | 3 | Describe the rationale for the review in the context of what is already known. Explain why the review questions/objectives lend themselves to a scoping review approach. | 3-9 |
| Objectives | 4 | Provide an explicit statement of the questions and objectives being addressed with reference to their key elements (e.g., population or participants, concepts, and context) or other relevant key elements used to conceptualize the review questions and/or objectives. | 9 |
| METHODS |  |  |  |
| Protocol and registration | 5 | Indicate whether a review protocol exists; state if and where it can be accessed (e.g., a Web address); and if available, provide registration information, including the registration number. | 10 |
| Eligibility criteria | 6 | Specify characteristics of the sources of evidence used as eligibility criteria (e.g., years considered, language, and publication status), and provide a rationale. | 10-12 |
| Information sources* | 7 | Describe all information sources in the search (e.g., databases with dates of coverage and contact with authors to identify additional sources), as well as the date the most recent search was executed. | 12-13 |
| Search | 8 | Present the full electronic search strategy for at least 1 database, including any limits used, such that it could be repeated. | 12 |
| Selection of sources of evidence† | 9 | State the process for selecting sources of evidence (i.e., screening and eligibility) included in the scoping review. | 12-13 |
| Data charting process‡ | 10 | Describe the methods of charting data from the included sources of evidence (e.g., calibrated forms or forms that have been tested by the team before their use, and whether data charting was done independently or in duplicate) and any processes for obtaining and confirming data from investigators. | 15 |
| Data items | 11 | List and define all variables for which data were sought and any assumptions and simplifications made. | 15 |
| Critical appraisal of individual sources of evidence§ | 12 | If done, provide a rationale for conducting a critical appraisal of included sources of evidence; describe the methods used and how this information was used in any data synthesis (if appropriate). | N/A |
| Synthesis of results | 13 | Describe the methods of handling and summarizing the data that were charted. | 13 |
| RESULTS |  |  |  |
| Selection of sources of evidence | 14 | Give numbers of sources of evidence screened, assessed for eligibility, and included in the review, with reasons for exclusions at each stage, ideally using a flow diagram. | 14 |
| Characteristics of sources of evidence | 15 | For each source of evidence, present characteristics for which data were charted and provide the citations. | 15-19 |
| Critical appraisal within sources of evidence | 16 | If done, present data on critical appraisal of included sources of evidence (see item 12). | N/A |
| Results of individual sources of evidence | 17 | For each included source of evidence, present the relevant data that were charted that relate to the review questions and objectives. | 15-19 |
| Synthesis of results | 18 | Summarize and/or present the charting results as they relate to the review questions and objectives. | 20-25 |
| DISCUSSION |  |  |  |
| Summary of evidence | 19 | Summarize the main results (including an overview of concepts, themes, and types of evidence available), link to the review questions and objectives, and consider the relevance to key groups. | 25-26 |
| Limitations | 20 | Discuss the limitations of the scoping review process. | 26-27 |
| Conclusions | 21 | Provide a general interpretation of the results with respect to the review questions and objectives, as well as potential implications and/or next steps. | 27-28 |
| FUNDING |  |  |  |
| Funding | 22 | Describe sources of funding for the included sources of evidence, as well as sources of funding for the scoping review. Describe the role of the funders of the scoping review. | N/A |

## S3 Appendix. Inclusion and Exclusion Criteria

| Participants |  |
| --- | --- |
| **Include:**<br>Participants aged ≥18 enrolled in higher education (undergraduate or postgraduate) (inclusion applies if at least one participant meets these criteria; number and characteristics of university student participants to be documented when reported). | **Exclude:**<br>Studies with no university student participants.<br>Studies that provide no participant information, including no demographic details and no indication of university student participation. |
| Intervention / Exposure |  |
| **Include:**<br>Interventions with secular aims (e.g., for health or well-being promotion).<br>Interventions incorporating Buddhist-derived practice elements other than mindfulness, or combining mindfulness with other Buddhist-derived practice elements.<br>Interventions grounded in identifiable Buddhist pedagogical frameworks, including but not limited to: Threefold Training (*tisikkhā*); Three Marks of Existence (*tilakkhaṇa*); Four Immeasurables (*brahmavihāras*); Tibetan mind training (*lojong*)<br>Must include ethics (*sīla*) and/or insight/wisdom (*vipassanā/paññā*) aspects of the threefold training (*tisikkhā*). | **Exclude:**<br>Interventions with religious or soteriological aims explicitly oriented toward Buddhist spiritual goals, such as favourable rebirth or spiritual enlightenment (*nibbāna*).<br>Buddhist-derived practice elements incorporated without grounding within a Buddhist pedagogical framework (e.g. mindfulness meditation (MM), loving-kindness meditation (LKM), compassion meditation (CM) or appreciative joy meditation (AJM) without grounding within the threefold training (*tisikkhā*) or four immeasurables (*brahmavihāras*)). |
| Outcome |  |
| **Include:**<br>Any reported positive or adverse outcomes, whether from pilot/testing phases or full implementation, documented in either peer-reviewed or grey literature. | **Exclude:**<br>No reported outcomes available in either grey or peer reviewed literature. |
| Study Characteristics |  |
| **Include:**<br>Primary empirical intervention studies (e.g., RCTs, controlled trials, quasi-experimental studies, qualitative intervention evaluations, mixed-methods intervention studies). | **Exclude:**<br>Review papers (e.g. systematic reviews, scoping reviews, narrative reviews). |

## S4 Appendix. Electronic search strategy

### EMBASE, MEDLINE, PsychINFO, Global Health and AMED

#### Search One - Date searched 11.07.25

##### SEARCH TERMS

second-generation mindfulness-based intervention OR buddhist-derived intervention OR buddhist meditation practices OR sila OR samatha OR samadhi OR concentration meditation OR loving-kindness meditation OR compassion meditation OR vipassana OR panna OR prajna OR wisdom OR nonattachment OR attachment-based compassion therapy OR cognitively-based compassion training OR compassion cultivation training OR ethical mindfulness OR ethics-oriented mindfulness training OR four immeasurables program OR meditacion-fluir OR insight-based mindfulness program OR meditation awareness training OR mindfulness-based attention training OR mindfulness-based eating awareness training OR meditation-based lifestyle modification OR mindfulness-based positive psychology OR mindful attention training OR mindfulness-integrated cognitive behavior therapy OR mindful lovingkindness-compassion program OR psychoneuroendocrinoimmunology-based meditation OR sustainable compassion training

AND

students OR university students OR college students

Limits: 2010-current

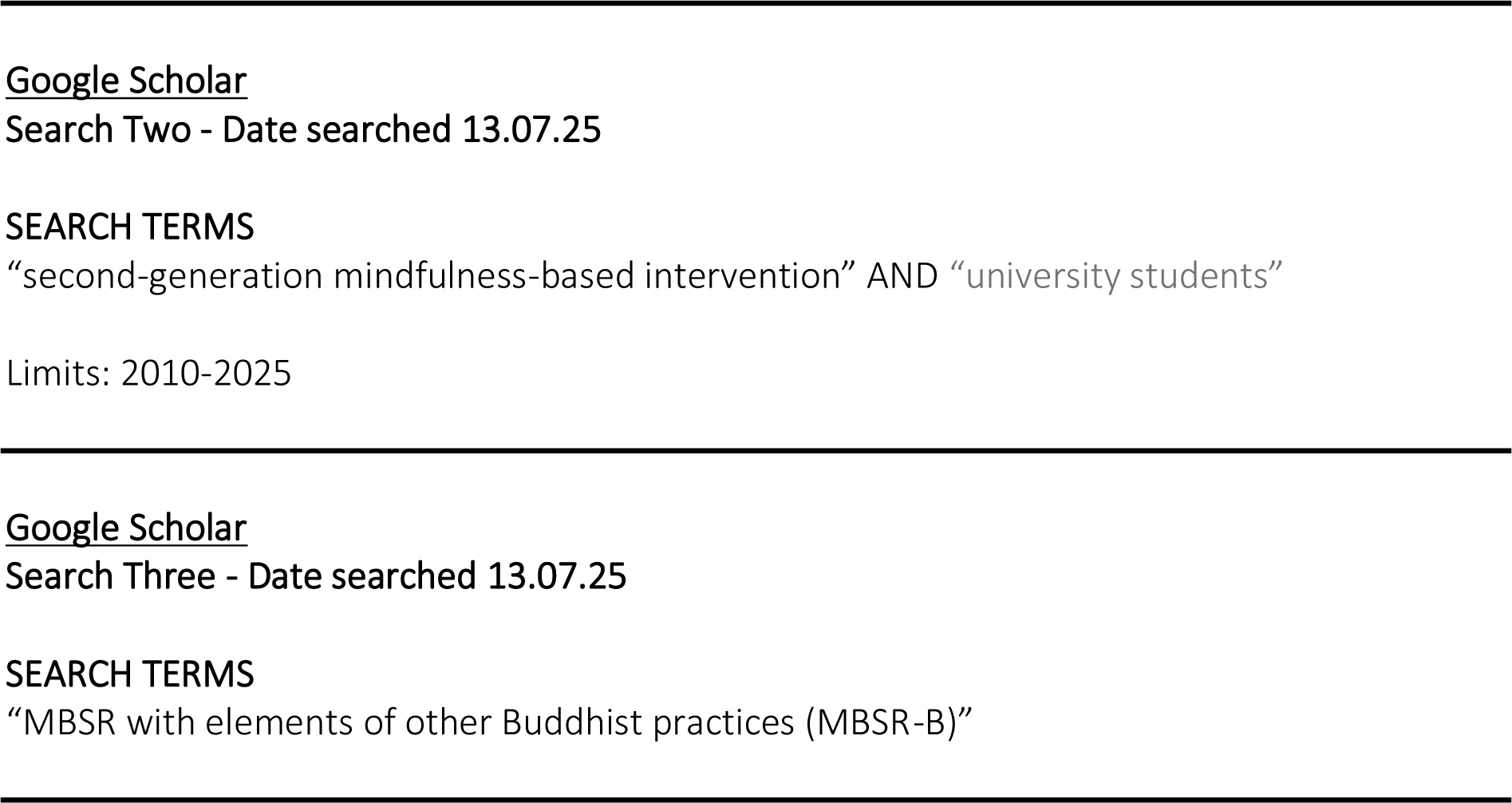
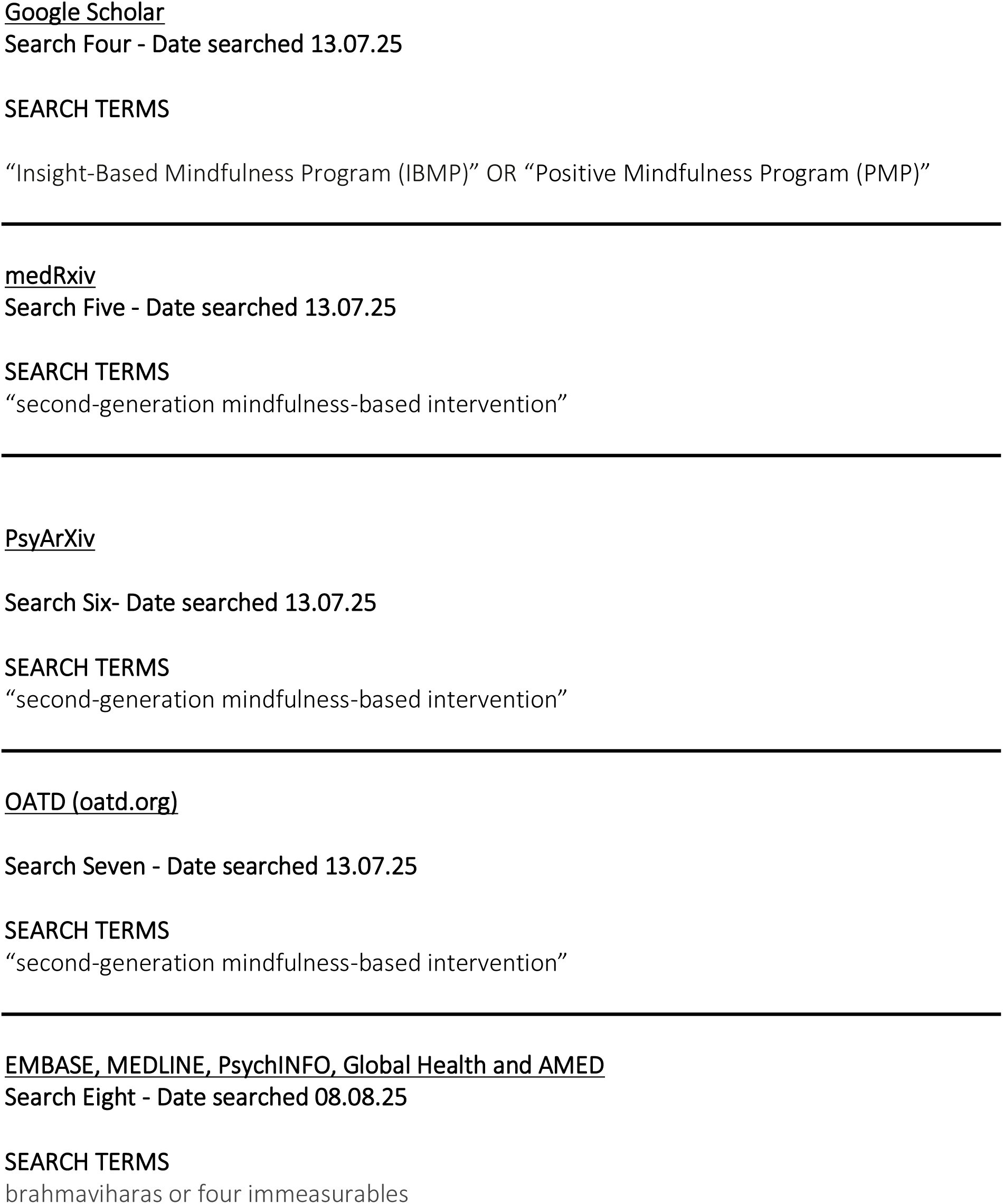

## S5 Appendix. SG-MBIs for university students

### Cognitively-Based Compassion Training (CBCT) (138)

CBCT was developed in 2005 at Emory University by Dr. Lobsang Tenzin Negi. Whilst delivered entirely in secular terms, it is based on the Indo-Tibetan Buddhist practices of mind training (*lojong*) and pedagogical framework of the ‘stages of the path for spiritual development’ (*lamrim*) (200). In relation to ethics/behaviour-change (*sīla*) it includes developing appreciation, gratitude, affection, empathy and realizing ‘active compassion for others’. In relation to concentration and tranquillity (*samādhi*) it includes ‘…*shamatha* and *vipassana* to calm and focus the mind…’ and a focus on developing ‘…attention and stability of mind’, and in relation to wisdom (*paññā*) it includes ‘…cultivating insight into the nature of mental experience’ (138).

### Compassion Cultivation Training (CTT) (124,142,143,165)

CCT was developed by a multidisciplinary group at Stanford University’s Center for Compassion and Altruism Research and Education (CCARE). This group included Thupten Jinpa, Ph.D., a scholar (*dge shes*) trained in the Tibetan Gelug (*dge lugs*) tradition. Practice elements within CCT include loving-kindness meditation (*metta*) and giving and receiving mediation (*tonglen*) which: ‘…have been secularized in the context of CCT but have been practiced by Buddhist practitioners for over 2,000 years’ (124). In a comparative analysis of compassion cultivation in Indo-Tibetan Mahāyāna Buddhist contexts and in CCT, it was concluded that ‘…more than the mere technique of tonglen, CCT pedagogy is infused with Buddhist ideas at every step of the program’ (201). In relation to ethics/behaviour-change (*sīla*) it includes exercises directed towards developing loving kindness and compassion for others, and embracing shared common humanity. In relation to concentration and tranquillity (*samādhi*) it includes basic skills to still and focus the mind through breath focused meditation, and in relation to wisdom (*paññā*) it includes developing an appreciation of how human beings are deeply interconnected.

### Mindfulness with additional ethical instruction (EthicalM) (22)

Chen and Jordan (2020) researched the effects of practicing mindfulness with additional ethical instruction (EthicalM) on well-being and prosocial behaviour (22). EthicalM was identified by the reviewers as encompassing the Buddhist pedagogical framework of the threefold training (*tisikkhā*). In relation to ethics/behaviour-change (*sīla*) participants were encouraged to reflect on their connection with all living beings and the importance of respecting and not harming them. In relation to concentration and tranquillity (*samādhi*) the intervention included breath-focused meditation and awareness of bodily sensations, emotions, and thoughts, and in relation to wisdom (*paññā*), EthicalM included guidance on recognising “wholesome” and “unwholesome” mental states, interconnectedness and the temporary nature of emotions and thoughts.

### Flow Meditation (FM) (162)

The purpose of Flow Meditation (FM; *Meditación-Fluir*) (202) is ‘…for participants to learn to allow their thoughts to flow, without trying to modify them or interfere with them’ (162). As well as aspects from Zen and ‘*Vipassana* meditation’ this intervention included exercises from MBSR, Acceptance and Commitment Therapy (ACT) (203) and logotherapy (204). In relation to ethics/behaviour-change (*sīla*) it includes ethical aspects and values. In relation to concentration and tranquillity (*samādhi*) it includes learning how to live in the present moment with awareness, and in relation to wisdom (*paññā*), it includes developing awareness of the impermanency of thoughts and of everything (*anicca*). On the basis of these aspects the reviewers agreed that the intervention could be considered as encompassing the Buddhist pedagogical framework of the threefold training (*tisikkhā*).

### Insight-Based Mindfulness Program (IBMP) (129)

The Insight-Based Mindfulness Program (IBMP) was developed by Dr. Somboon Jarukasemthawee, who is currently based at Chulalongkorn University, Thailand. IBMP was based on the Buddhist pedagogical framework of the three marks of existence (*tilakkhaṇa*). It incorporates directed mindfulness and psychoeducation to promote a secular-based insight into the three marks. Although IBMP does not include an explicit ethics/behaviour-change (*sīla*) component, in relation to concentration and tranquillity (*samādhi*) it includes a ‘mindfulness of concentration’ session and in relation to wisdom (*paññā*) it includes an insight focused component.

### Meditation Awareness Training (MAT) (159,160)

Meditation Awareness Training (MAT) was developed in the UK. It has a basis in the Buddhist pedagogical framework of the threefold training (*tisikkhā*), and is ‘…firmly grounded in Buddhist principles’; however, it: ‘…makes no explicit reference to Buddhist terms’ (159). In relation to ethics/behaviour-change (*sīla*) MAT includes elements designed to foster improved self-control and ethical awareness, patience, generosity, and compassion. In relation to concentration and tranquillity (*samādhi*) it includes meditation and mindfulness, and in relation to wisdom (*paññā*) insight meditation techniques are integrated ‘…to encourage a gradual familiarization with (and ideally a preliminary realization of) concepts such as impermanence and emptiness’ (159).

### Mindfulness-Based Intervention including Concentration- and Ethics-based Practices (MBI-CE) and Mindfulness-Based Intervention including Concentration-, Ethics- and Wisdom-based Practices (MBI-CEW) (119)

To examine the impact on prosocial behaviour of incorporating Buddhist ethics-based practices versus Buddhist ethics- and wisdom-based practices, Furnell, Van Gordon and Elander (119) based at the University of Derby, UK, developed Mindfulness-Based Intervention including concentration- and ethics-based practices (MBI-CE) and a MBI including concentration-, ethics-, and wisdom-based practices (MBI-CEW). MBI-CE applied the Buddhist pedagogical framework of the four immeasurables (*brahmavihāras*) and MBI-CEW applied the Buddhist pedagogical frameworks of both the threefold training (*tisikkhā*) and the three marks of existence (*tilakkhaṇa*). In relation to ethics/behaviour-change (*sīla*) both MBI-CE and MBI-CEW included explorations of ethics and on awareness of feelings and learning to respond rather than react. Whereas MBI-CE also included ethics-based practices which ‘…mainly consisted of the Four Immeasurables (*Brahmavihārās*), including loving-kindness, compassion, empathetic-joy, and equanimity meditation practices’ (119). In relation to concentration and tranquillity (*samādhi*) both MBI-CE and MBI-CEW included mindfulness of the breath and body scan exercises as practices towards present-centred awareness, and in relation to wisdom (*paññā*), only MBI-CEW included ‘wisdom-based practices’ which included contemplations on impermanence (*anicca*), non-attachment, interdependence and no-self (*anatta*).

### Mindfulness-based positive psychology (MBPP) (167,168)

Mindfulness-based positive psychology (MBPP) is an intervention integrating positive psychology with an SG-MBI. It was developed by Xianglong Zeng at the Faculty of Psychology, Beijing Normal University in China. The Buddhist pedagogical framework of the four immeasurables (*brahmavihāras*) formed the basis for meditation techniques within the intervention. In relation to ethics/behaviour-change (*sīla*) MBPP includes the four immeasurables (*brahmavihārās*) which can be applied as a basis for ethical decision making and in relation to concentration and tranquillity (*samādhi*) mindfulness meditation (body scanning) is included in MBPP; however, wisdom-based practices (*paññā*) are not included within MBPP.

### MBSR with elements of other Buddhist practices (MBSR-B) (42,43)

MBSR-B is a modification of mindfulness-based stress reduction (MBSR) (23–25) which includes additional modules based on other Buddhist practices. MBSR-B was developed in Switzerland for the purpose of assessing the stress-attenuating effects of MBSR) and MBSR with elements of other Buddhist practices (MBSR-B). MBSR-B applies two Buddhist pedagogical frameworks; the threefold training (*tisikkhā*) and the three marks of existence (*tilakkhaṇa*). In relation to ethics/behaviour-change (*sīla*) informal practices within MBSR-B included, for example, abstaining from one minor unethical action for the week (like gossiping), practicing one generous action, using informal loving-kindness intentions during difficult conversations and using informal compassionate intentions when meeting someone who faces difficulties. In relation to concentration and tranquillity (*samādhi*) the MBSR-B program followed the same outline as the standard MBSR, and in relation to wisdom (*paññā*), MBSR-B included being aware of the impermanence (*anicca*) of emotions, and of moments of “selfing” verses moments of mindful activities.

### Three Marks Intervention (TMI) (133)

The three marks intervention was a video featuring Robert J. Klein (133) discussing the three marks concepts (impermanence [*anicca*], non-self [*anatta*] and unsatisfactoriness [*dukkha*]). Klein has studied Buddhist thought for 20 years and has an extensive background in meditation and Buddhist retreats and the video was based on texts approved by a prominent, well-published Buddhist scholar and teacher. Whilst centred on the Buddhist pedagogical framework of the three marks of existence (*tilakkhaṇa*), insight into which is characteristic of Buddhist wisdom (*paññā*), ethics/behaviour-change (*sīla*) and concentration and tranquillity (*samādhi*) were not included within this experimental study.

## S6 Appendix. Outcome Measures and Well-being Domains Addressed in SG-MBIs for University Students

| SG-MBI | Physical well-being | Mental well-being | Social well-being | Spiritual well-being |
| --- | --- | --- | --- | --- |
| CBCT | • Exercise (2Q)<br>• PSQI<br>• SUI | • DASS-21<br>• R-UCLA |  | • CLHS |
| CCT |  | • BRS<br>• CESD-R<br>• DASS-21<br>• DERS<br>• FFMQ-SF<br>• JSPE-S<br>• KIMS<br>• MBI-SS<br>• MHI-5<br>• PHI | • IRI | • CSP<br>• SCS<br>• SCS-SF |
| EthicalM |  | • BDI-II<br>• CSIG<br>• MAAS<br>• PSS<br>• PWB<br>• SDS<br>• SHS<br>• SWLS | • IRI |  |
| FM | • HLQ<br>• LQ |  |  |  |
| IBMP |  | • DASS-21 (Thai)<br>• EWBS<br>• FMI |  | • IS |
| MAT |  | • DASS-21<br>• MAAS<br>• PANAS |  |  |
| MBI-CE /<br>MBI-CEW |  | • MAAS | • PSA<br>• RQ<br>• WCS |  |
| MBPP |  | • DASS-21<br>• mDES<br>• PHLMS<br>• PWB<br>• SWLS |  | • AJS<br>• GRAT<br>• ML<br>• SCS-SF<br>• SOFI |
| MBSR-B | • ANS<br>• CO<br>• HPA<br>• SAM<br>• RR<br>• TPR | • SRA<br>• PASA |  |  |
| TMI |  | • DART<br>• PASA<br>• STAI-S |  | • NAS |

### Outcome Measure Abbreviations Table

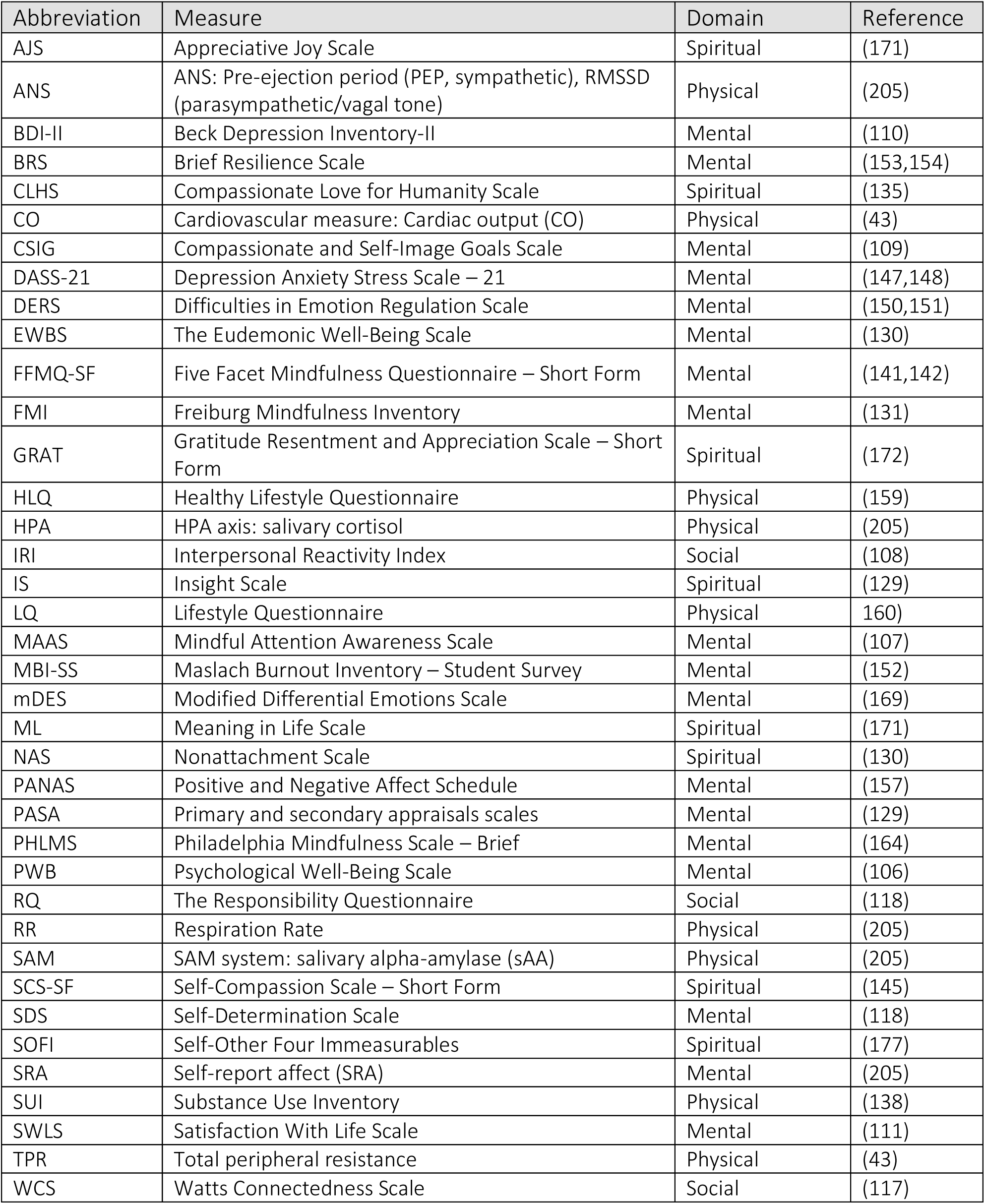

## Notes

### Competing Interest Statement

The authors have declared no competing interest.

### Funding Statement

The authors received no specific funding for this work.

### Author Declarations

This study is a scoping review of previously published literature and did not involve primary data collection from human participants. As such, it did not require institutional review board (IRB) approval or exemption.

